# Mathematical Modeling and Optimal Control Analysis of COVID-19 in Ethiopia

**DOI:** 10.1101/2020.07.23.20160473

**Authors:** Haileyesus Tessema Alemneh, Getachew Teshome Tilahun

## Abstract

In this paper we developed a deterministic mathematical model of the pandemic COVID-19 transmission in Ethiopia, which allows transmission by exposed humans. We proposed an SEIR model using system of ordinary differential equations. First the major qualitative analysis, like the disease free equilibruim point, endemic equilibruim point, basic reproduction number, stability analysis of equilibrium points and sensitivity analysis was rigorously analysed. Second, we introduced time dependent controls to the basic model and extended to an optimal control model of the disease. We then analysed using Pontryagin’s Maximum Principle to derive necessary conditions for the optimal control of the pandemic. The numerical simulation indicated that, an integrated strategy effective in controling the epidemic and the gvernment must apply all control strategies in combating COVID-19 at short period of time.

## 1 Introduction

Corona virus disease (COVID-19) which is caused by novel corona virus is a respiratory illness that can spread from person to person [1, 2, 3]. People with COVID-19 have had a wide range of symptoms reported – ranging from mild symptoms to severe illness. Symptoms may appear 2-14 days after exposure to the virus. People with these symptoms may have COVID-19: fever or chills, cough, shortness of breath or difficulty breathing, fatigue, muscle or body aches, headache, new loss of taste or smell, sore throat, congestion or runny nose, nausea or vomiting, diarrhea [1, 4]. The outbreak was first identified in Wuhan, China, in December, 2019, which has been spreading worldwide [5, 6]. Following the day of the outbreak of the pandemic to present more than seven million globally confirmed cases and half million deaths as well as four million recovered numbers [7]. In Africa also the virus spread to 57 countries [1, 7]. Through out Africa up to Jun 14, 2020 confirmed cases are 235,707, number of deaths are 6,283 and recoveries are 36,850 [8].

In order to combat this pandemic, different preventive measures are recommended, such as avoiding close contact with sick people, avoiding touching the eyes, nose and mouth with unwashed hands, washing hands often with soap and water for at least 20 seconds, using an alcohol-based hand sanitizer containing at least 60% alcohol when soap and water are not available [1, 3, 4]. Limit the number of people you have close contact with, or are visited by, to a few at a time. For infected individuals, staying home, covering cough or sneeze with a tissue, then throw the tissue in the trash, and clean and disinfect frequently touched objects are recommended for controlling COVID-19, perform home quarantine for 14 days after the last contact with a patient with confirmed COVID-19 [9, 10].

Mathematical models with optimal control analysis has become an important tool in order to understand the dynamics of disease transmission and decision making processes regarding intervention programs for the disease control. COVID-19 have been modeled by very few researchs after the outbreak of the disease. The study by [11], used an SIR model to predict the magnitude of the COVID-19 epidemic in Pakistan and compared the numbers with the reported cases on the national database. They predicted that 90% of the population will have become infected with the virus if policy interventions seeking to curb this infection are not adopted aggressively. The study presented in [12] used SEIR compartments by considering limited parameters, from January 22, 2020 to March 3, 2020 and Prediction SEIR forecasted by using SEIR model They also looked at the feelings, current disease trends and economic and political impacts. The article [13], proposed conceptual models for the COVID-19 outbreak in Wuhan with the consideration of individual behavioural reaction and governmental actions, e.g., holiday extension, travel restriction, hospitalisation and quarantine. The article published by [12] also proposed SEIR model by incorporating the intrinsic impact of hidden exposed and infectious cases on the entire procedure of epidemic, which is difficult for traditional statistics analysis. The other study is done in China using SIR by considering logistic growth [11]. Using their model, the study approximated future number of cases in china and proposed possible controlling mechanisms. The study [14] simulated the ongoing trajectory of Covid 19 outbreak in Wuhan using an age-structured SEIR model for several physical distancing measures. The article presented by [15], proposed a compartmental model by dividing the quarantined individuals in to two sub-groups. They developed an SEIRU model where R and U are quarantine-infected individuals expected to recover and meet undetectable criteria. A more detail model is proposed by [16], and studied a mathematical modeling of the spread of the COVID-19 taking into account the undetected infections, in China. The study in [17], proposed a new epidemic model in the case of Italy that discriminates between infected individuals depending on whether they have been diagnosed and on the severity of their symptoms. However, all of the above models studied the dynamics of disease transmission, they did not study finding an optimal control strategy using the maximum princeiple of Pontryagn.

From all the above studies we can understand that, COVID-19 have been modeled by considering different situations. In the case of Ethiopia, the proposed model can’t reflect the real situation of the country. Currently Ethiopia is implementing different measure to control COVID-19 including implementing state of emergency, counry border closing and mandatory quarantine of new arrivals for 14 days. herefore, in the present paper we extend the SEIR model with the aforementioned control measures and propose an optimal control strategy.

The paper is organized as follows. In Section 2 was devoted to the model description and the underlying assumptions. In Section 3 we carry out mathematical analysis of this COVID-19 model. In Section 4 we propose an optimal control problem and and the optimal control analysis are presented. In Section 5 numerical simulation and calibration of the model was implemented for the various strategies considered in this work. The conclusion was presented in Section 6.

## 2 Model Description and Formulation

In this model the entire population is divided into four sub-populations: Susceptible individuals (denoted by *S*) are those who are not infected by the disease pathogen but there is a possibility to be infected. Exposed individuals (denoted by *E*) are individuals who are in the incubation period after being infected and have no visible clinical signsand these individuals could infect other people with a higher probability than people in the infected compartments. After the incubation period, the person passes to the infected compartment; infected individuals (denoted by *I*) are individuals who developed the symptom of the disease. Recovered individuals (denoted by *R*)are those individuals who recovered from the disease.

Individuals are recruitment at a rate *π* is either through immigration or birth. The susceptible individuals got infection of COVD-19 disease at a contact rate of *β* either from infected with probability of *σ*_1_ or from exposed individuals with probability of *σ*_2_ and move to the exposed compartment. The exposed individuals become infectious and join the infected compartment at *τδ* and the remaining proportion of this exposed individuals develop natural immunity and recovered from the disease. By the treatment given, the infected individuals will recover and move to the recovered compartment at a rate of *E*, or may die due to the disease at a rate of *ρ*. The recovered individuals become again susceptible to the disease at a rate of *η*. The whole population have an average death rate of *µ*. Table 1 shows the description of model parameters. The flow diagram of the model is shown in Figure 1 below.

**Table 1:** Description of parameters of the model (1).

| Parameter | Description |
| --- | --- |
| $\pi$ | Ricurement rate of individuals |
| $\beta$ | Contact rate of susceptible individuals |
| $\rho$ | Death rate due to disease |
| $\delta$ | Proportion of exposed individuals leaving the compartment. |
| $\varepsilon$ | Recovery rate of individuals from the disease |
| $\tau$ | Proportion of exposed individuals who join infected compartment |
| $\mu$ | Natural death rate |
| $\eta$ | Proportion of recovered individuals to be susceptible. |

**Figure 1:**
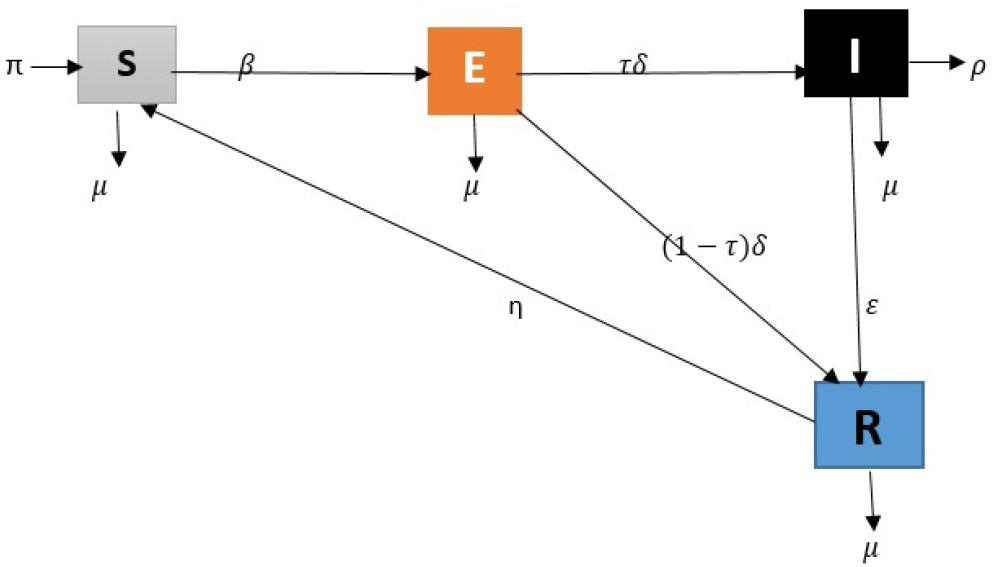
Compartmental flow diagram of the pandemic COVID 19 transmission

With regards to the above asumptions, the model is governed by the following system of differential equation:

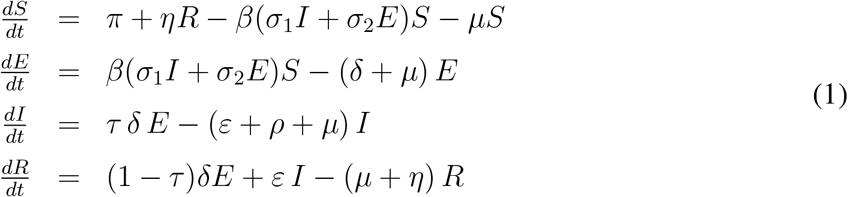

With the initial condition

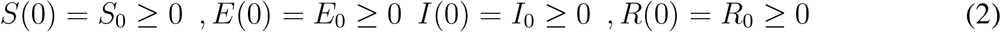

## 3 Model Analysis

### 3.1 Invariant Region

Let us determine a region in which the solution of model (1) is bounded. For this model the total population is *N* (*S, E, I, R*) = *S*(*t*) + *E*(*t*) + *I*(*t*) + *R*(*t*). Then, differentiating *N* with respect to time we obtain:

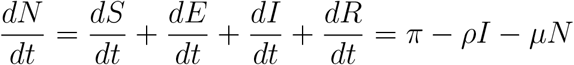

If there is no death due to the disease, we get

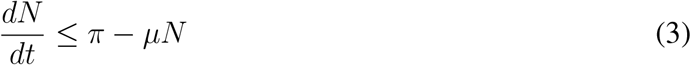

After solving equation (3) and evaluating it as *t → ∞*, we got

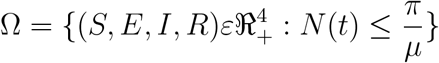

Which is the feasible solution set for the model (1) and all the solution set is bounded in it.

### 3.2 Positivity of Solutions

#### Theorem 3.1.

*If S*(0) > 0, *E*(0) > 0, *I*(0) > 0, *R*(0) > 0 *are positive in the feasible set* Ω, *then the solution set* (*S*(*t*), *E*(*t*), *I*(*t*), *R*(*t*)) *of system (1) is positive for all t ≥* 0.

*Proof*. : From the first equation of the system

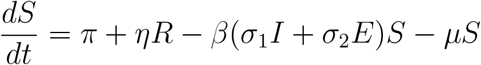

which can be taken as

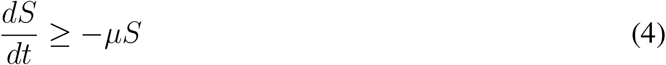

After evaluating equation (4), we obtain

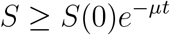

Similarly, we obtain

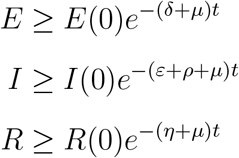

Therefore, all the solution sets are positive for *t ≥* 0.

### 3.3 Disease Free Equilibrium Point(DFEP)

When there is no infectious person of the disease in the population, I.e *E* = *I* = 0, the disease free equilibrium occur and is obtained by taking the right side of Eq. (1) equal to zero. Therefore the disease free equilibrium point is given by:

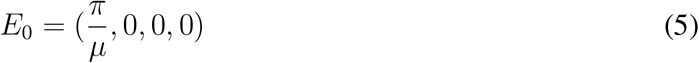

### 3.4 Basic reproduction number

We calculate the basic reproduction number _0_ of the system by applying the next generation matrix method as laid out by [18]. The first step is rewrite the model equations, starting with newly infective classes:

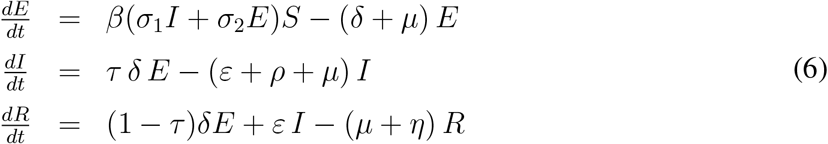

Then by the principle of next-generation matrix, the Jacobian matrices at DFE is given by

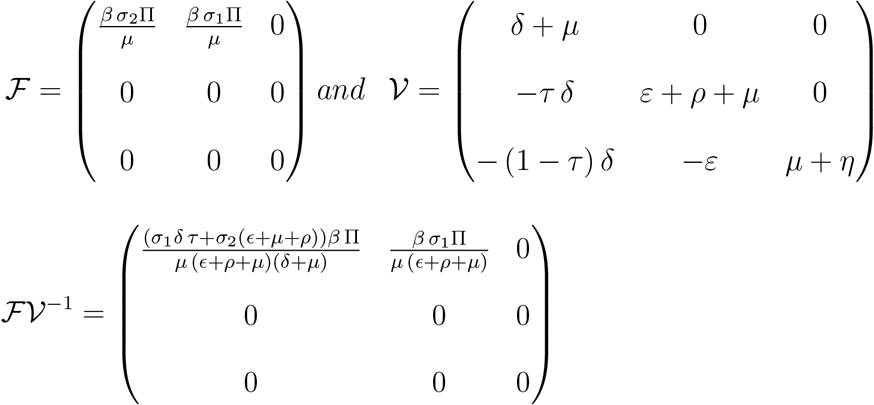

Therefore, the basic reproduction number is given us

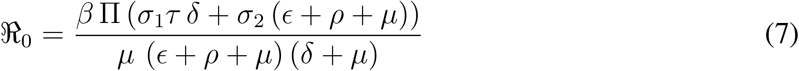

ℜ_0_ is a threshold parameter that represents the average number of infection caused by one infectious individual when introduced in the susceptible population [18].

### 3.5 Local Stability of DFEP

#### Theorem 3.2.

*The DFEP point is locally asymptotically stable if* ℜ_0_ < 1 *and unstable if* ℜ_0_ > 1. *Proof*. The Jacobian matrix, evaluated at the disease-free equilibrium *E*_0_, we get:

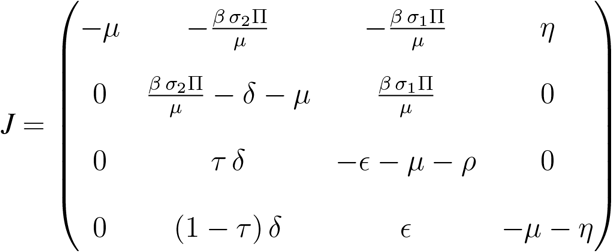

The characteristic polynomial from the Jacobian matrix is

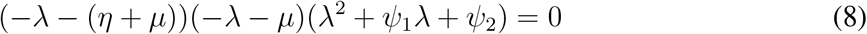

Where

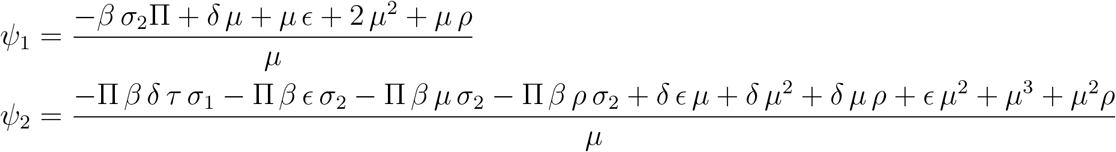

From the Eq.(8), we see that

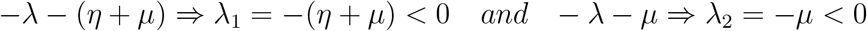

From the last expression, that is

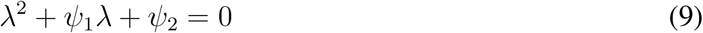

We applied Routh-Hurwitz criteria and by the principle Eq.(8) has strictly negative real root iff *ψ*_1_ > 0, *ψ*_2_ > 0 and *ψ*_1_*ψ*_2_ > 0. Clearly we see that *ψ*_1_ > 0 because it is the sum of positive parameters.

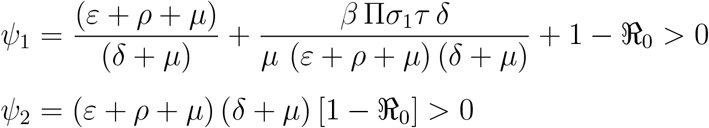

Hence the DFEP is locally asymptotically stable if ℜ_0_ < 1.□

### 3.6 Global Stability of DFEP

#### Theorem 3.3.

*The DEFP E*_0_ *of the model (1) is globally asymptotically stable if* ℜ_0_ < 1.

*Proof*. Consider the following Lyapunov function

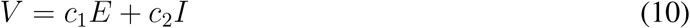

Differentiating equation (10) with respect to *t* gives

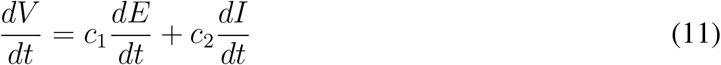

Substituting 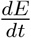 and 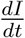 from the model (1), we get:

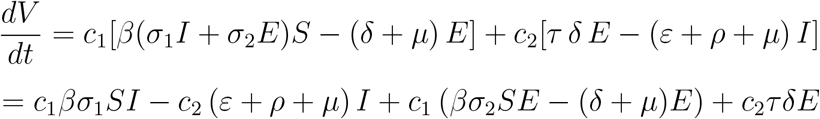

Here take 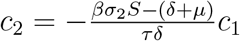, then we have

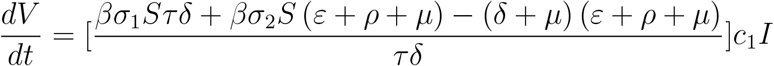

Taking *c*_2_ = 1, and substituting ℜ_0_ we get

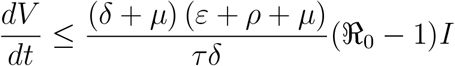

for 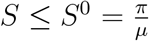 and 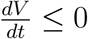 for ℜ_0_ < 1 and 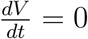 if and only if *I* = 0. This implies that the only trajectory of the system (1) on which 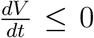 is *E*^0^. Therefore by Lasalle’s invariance principle, *E*^0^ is globally asymptotically stable in Ω.□

### 3.7 Existence of endemic equilibrium (EEP)

In the presence of disease in the population,(*S*(*t*) *≥* 0; *E*(*t*) *≥* 0; *I*(*t*) *≥* 0, *R*(*t*) *≥* 0), there exist an equilibrium point called endemic equilibrium point denoted by *E*^*\**^ = (*S*^*\**^, *E*^*\**^, *I*^*\**^, *R*^*\**^) *≠* 0.

#### Lemma 3.4.

*The Zika-only model has a unique endemic equilibrium if and only if* ℜ_0_ > 1.

*Proof*. It can be obtained by equating each equation of the model equal to zero. i.e

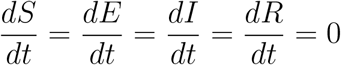

Then we obtain

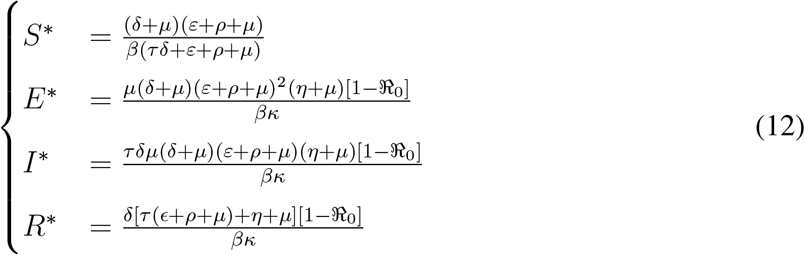

Where

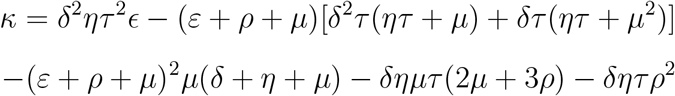

□

#### Theorem 3.5.

*The EEP is locally asymptotically stable if* ℜ_0_ > 1 *and unstable if otherwise. Proof*. The Jacobian matrix of system (1) at the DFEP is given by

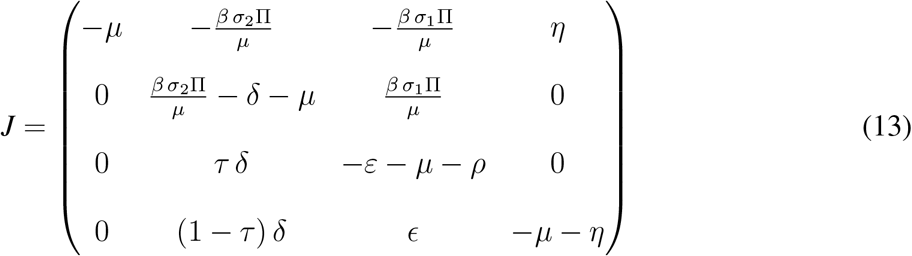

To determine the local stablity of endemic equilibrium, we used the center manifold theory [19], by taking *β* as a bufiracation parameter. let *S* = *z*_1_, *E* = *z*_2_, *I* = *z*_3_ and *R* = *z*_4_. In addition, using vector notation *z* = (*z*_1_, *z*_2_, *z*_3_, *z*_4_)^*T*^, formulated as 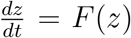, with *F* = (*f*_1_, *f*_2_, *f*_3_, *f*_4_, *f*_5_)^*T*^ and the disease free equilibrium is given by 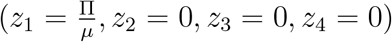. Then the value of *β* as a bifurcation parameter and solve ℜ_0_ = 1, which leads to

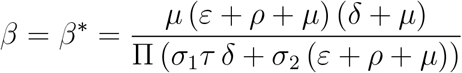

The right eigenvector, *w* = (*w*_1_, *w*_2_, *w*_3_, *w*_4_)^*T*^, associated with this simple zero eigenvalue can be obtained as:

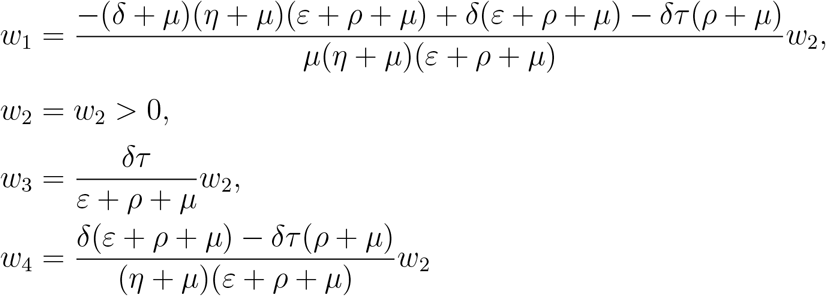

The left eigenvector, *v* = (*v*_1_, *v*_2_, *v*_3_, *v*_4_, *v*_5_), associated with this simple zero eigenvalue is given by

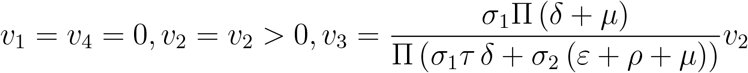

Since the first and fourth component of *v* are zero, we don’t need the derivatives of *f*_1_ and *f*_4_. From the derivatives of *f*_2_ and *f*_3_, the only ones that are nonzero are the following:

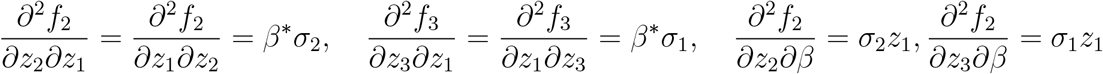

and all the other partial derivatives of are zero. The direction of the bifurcation at ℜ_0*d*_ = 1 is determined by the signs of the bifurcation coefficients *a* and *b*, obtained from the above partial derivatives, given respectively by

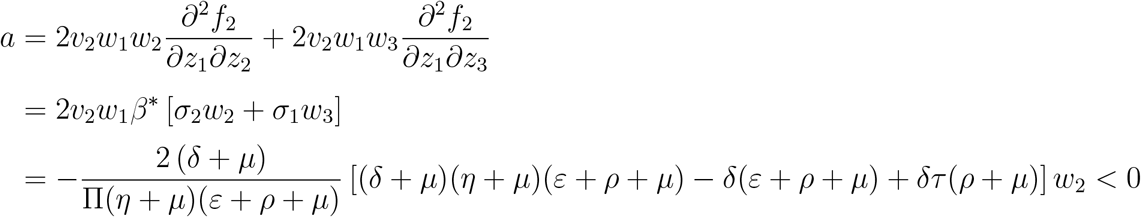

and

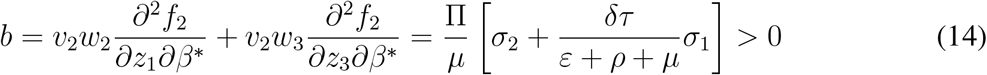

Do to the sign of the coefficient *a* < 0 and *b* > 0 at *β*^*\**^, by the theorem 4.1 stated in [19] system (11) undergo forward bifurcation at ℜ_0_ = 1 and the unique endemic equilibruim is locally assymptotically stable for ℜ_0_ > 1.□

### 3.8 Sensitivity Analysis

We carried out sensitivity analysis, on the basic parameters, to identify their effect to the transmition of the disease. To go through sensitivity analysis, we used the normalized sensitivity index definition as defined in [20]. The Normalized forward sensitivity index of a variable, ℜ_0_, that depends differentiably on a parameter, *p*, is defined as: 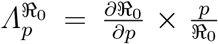 for *p* represents all the basic parameters. Here we have 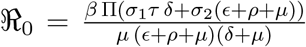. For the sensitivity index of ℜ_0_ to the parameters:

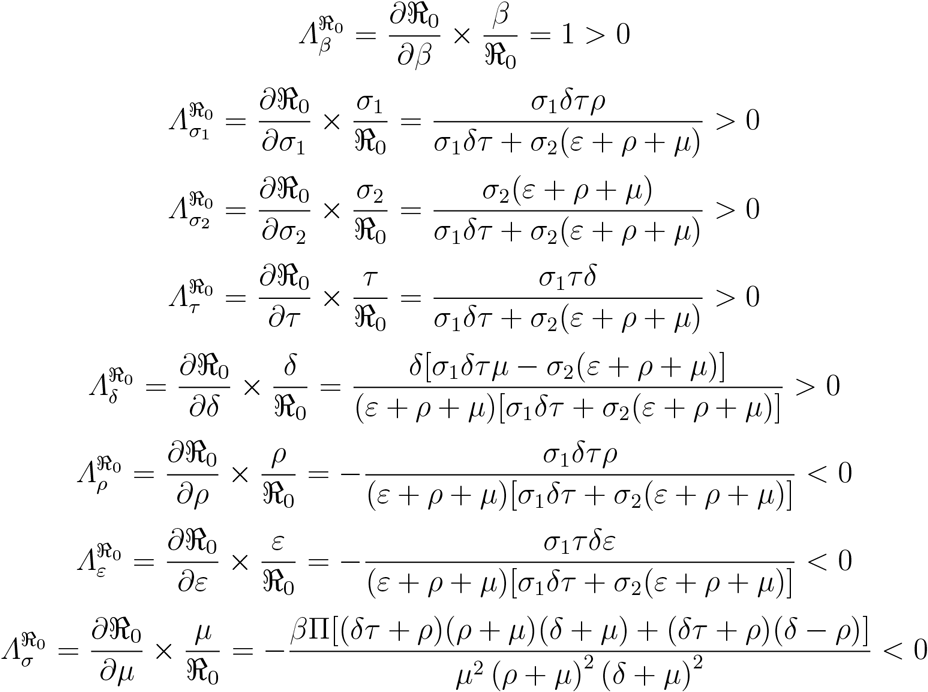

The sensitivity indices of the basic reproductive number with respect to main parameters are found in Table 2. Those parameters that have positive indices (Π, *β, σ*_1_, *σ*_1_, *and τ*) show that they have great impact on expanding the disease in the community if their values are increasing. Also those parameters in which their sensitivity indices are negative (*δ, ρ, ε, and µ*) have an effect of minimizing the burden of the disease in the community as their values increase. Therefore, research advice for stakholders to work on decreasing the positive indeces and increasing negative indices parameters

**Table 2:** Sensitivity indecies table.

| Parameter symbol | Sensitivity indecies |
| --- | --- |
| $\beta$ | +ve |
| $\sigma_1$ | +ve |
| $\sigma_2$ | +ve |
| $\tau$ | +ve |
| $\epsilon$ | -ve |
| $\delta$ | -ve |
| $\rho$ | -ve |
| $\mu$ | -ve |

## 4 Extension to an Optimal Control Model

Here we extend the basic model in (1) in to optimal control. As we indicated in section 1, Ethiopian goverment have started a lot of activities to combat COVID-19. Some of preventive activities are; forced isolation for 14 days for new arrivals, closing country border, quarantining and supporting infected individuals with medication, restricting number of sits by half in all transport system, convincing all media outlets to campaigning on COVID-19, announcing state of emergency and others. As we observe the started activities are not done in optimal level. Therefore, we want to show the concerned body the effectiveness of those activities if they are implemented in an optimal level. To perform optimal control, we categorize those activities in to three broad control strategies listed below.

i. All rounded prevention strategies including social distancing and personal hygiene (by this strategy we aimed to block susceptible from contacting the virus)
ii. Supporting infectives with medication (Optimal support of infected individual in quarantine center)
iii. Awareness creation through all Media outlets.

After incorporating the three controls in model (1) gives the following optimal control model in equation (15) below.

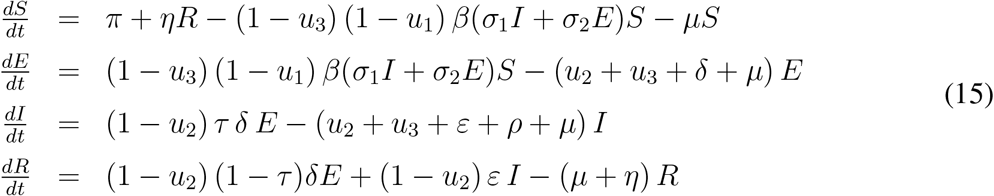

The purpose of introducing controls in the model is to find the optimal level of the intervention strategy required to reduce the spreads the pandemic in the population. Here we want to find the optimal values *u*_1_, *u*_2_ and *u*_3_ that minimizes the objective functional subject to the differential equations (15). The objective functional is given as

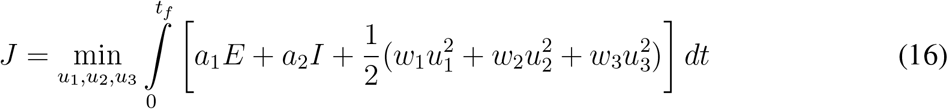

where *t*_*f*_ is the final time, *a*_1_ and *a*_2_ are weight costs of the Exposed humans and infected humans respectively while *w*_1_, *w*_2_ and *w*_3_ are weight costs for each individual control measure. In this paper, a quadratic function which satisfies the optimality conditions is considered for measuring the cost of the controls as applied by [21, 22, 23]. The goal is to find the optimal control 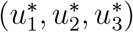 such that.

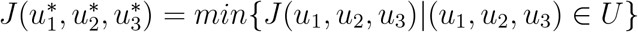

where the control set

*U* = *{*(*u*_1_, *u*_2_, *u*_3_) | *u*_*i*_(*t*) is lebesgue measurable on [0, *t*_*f*_], 0 *≤ u*_*i*_(*t*) *≤* 1, *i* = 1, 2, 3*}*

### 4.1 Hamiltonian and optimality equation

We used Pontryangin’s Maximum Principle [24] to drive the necessary conditions that an optimal control must satisfy. This principle converts equation (15) and (16) into a problem of minimizing point-wise Hamiltonian (*H*), with respect to *u*_1_(*t*), *u*_2_(*t*) *and u*_3_(*t*) as:

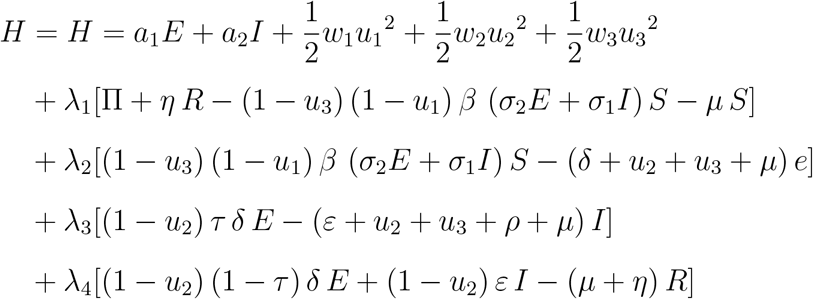

Where *λ*_*i*_, *i* = 1, 2, 3, 4 are the adjoint variable associated with *S, E, I, and R* to be determined suitably by applying Pontryagin’s maximal principle [24] and also using [25], the existence of an optimal control is guaranteed.

#### Theorem 4.1.

*For an optimal control set u*_1_, *u*_2_, *u*_3_ *that minimizes J over U, there are adjoint variables, λ*_1_, …, *λ*_4_ *such that:*

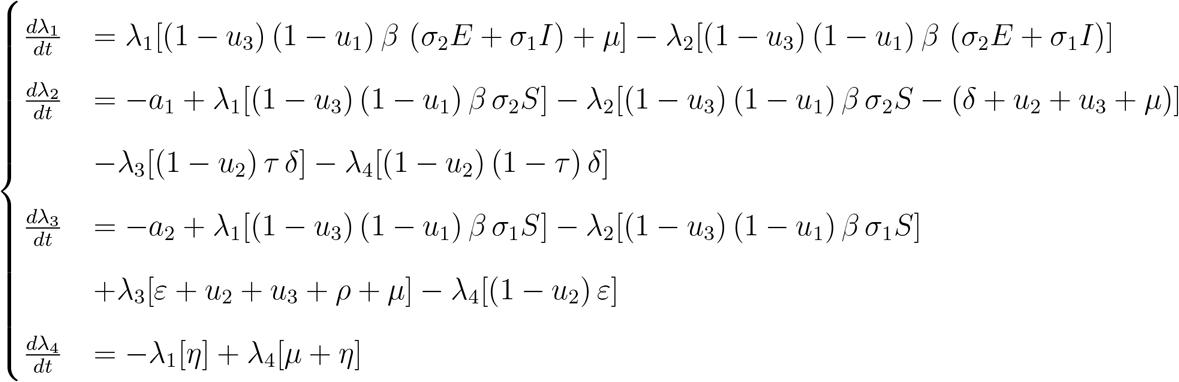

*With transversality conditions, λ*_*i*_(*t*_*f*_) = 0, *i* = 1, …, 4. *Furthermore, we obtain the control set* 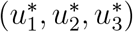 *characterized by*

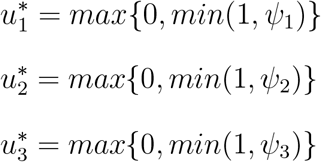

*Where*

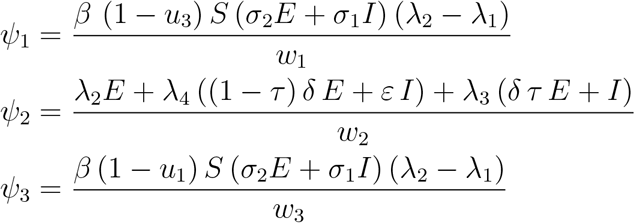

*Proof*. The adjoint equation and transversality conditions are standard results from Pontryagin’s maximum principle [24]. We differentiate Hamiltonian with respect to states *S, E, I* and *R* respectively, and then the adjoint system is written as

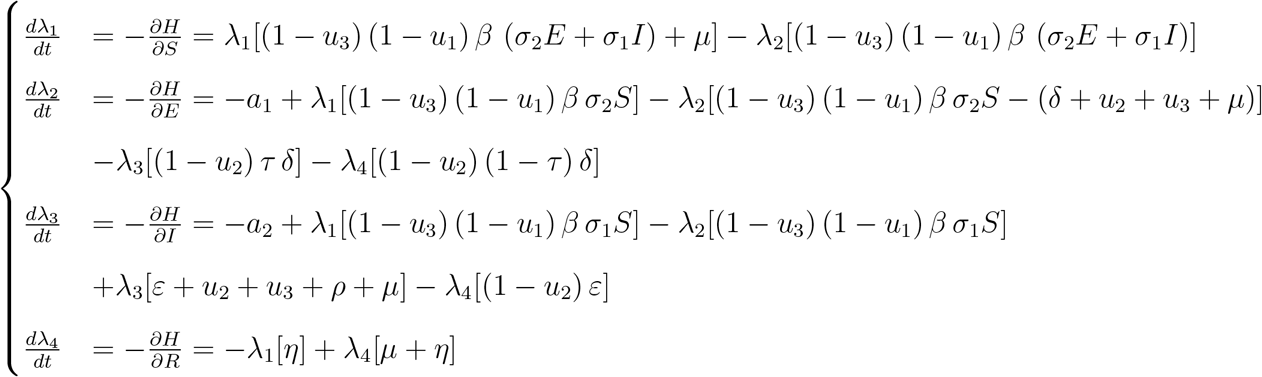

With transversality conditions, *λ*_*i*_(*t*_*f*_) = 0, *i* = 1, …, 4. Similarly by following the approach of Pontryagin et al. [24], the characterization of optimal controls 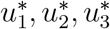, that is, the optimality equations are obtained based on the conditions: 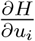, *for i* = 1, ‥, 3, which gives,

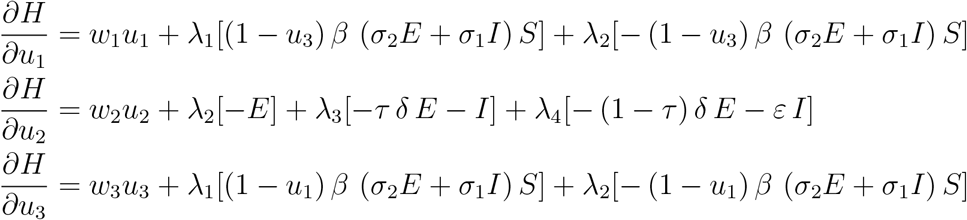

Setting 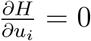 at 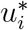, the results are

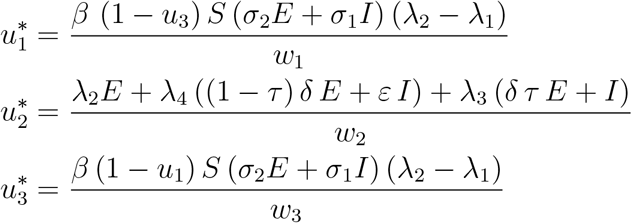

When we write by using standard control arguments involving the bounds on the controls, we conclude

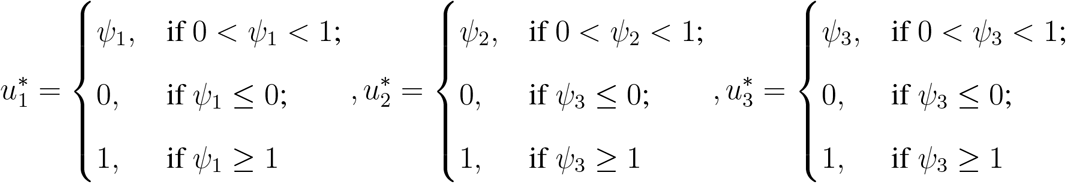

In compact notation

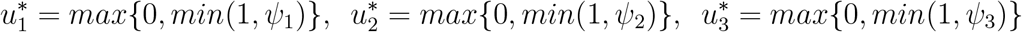

The optimality system is formed from the optimal control system (the state system) and the adjoint variable system by incorporating the characterized control set and initial and transversal condition

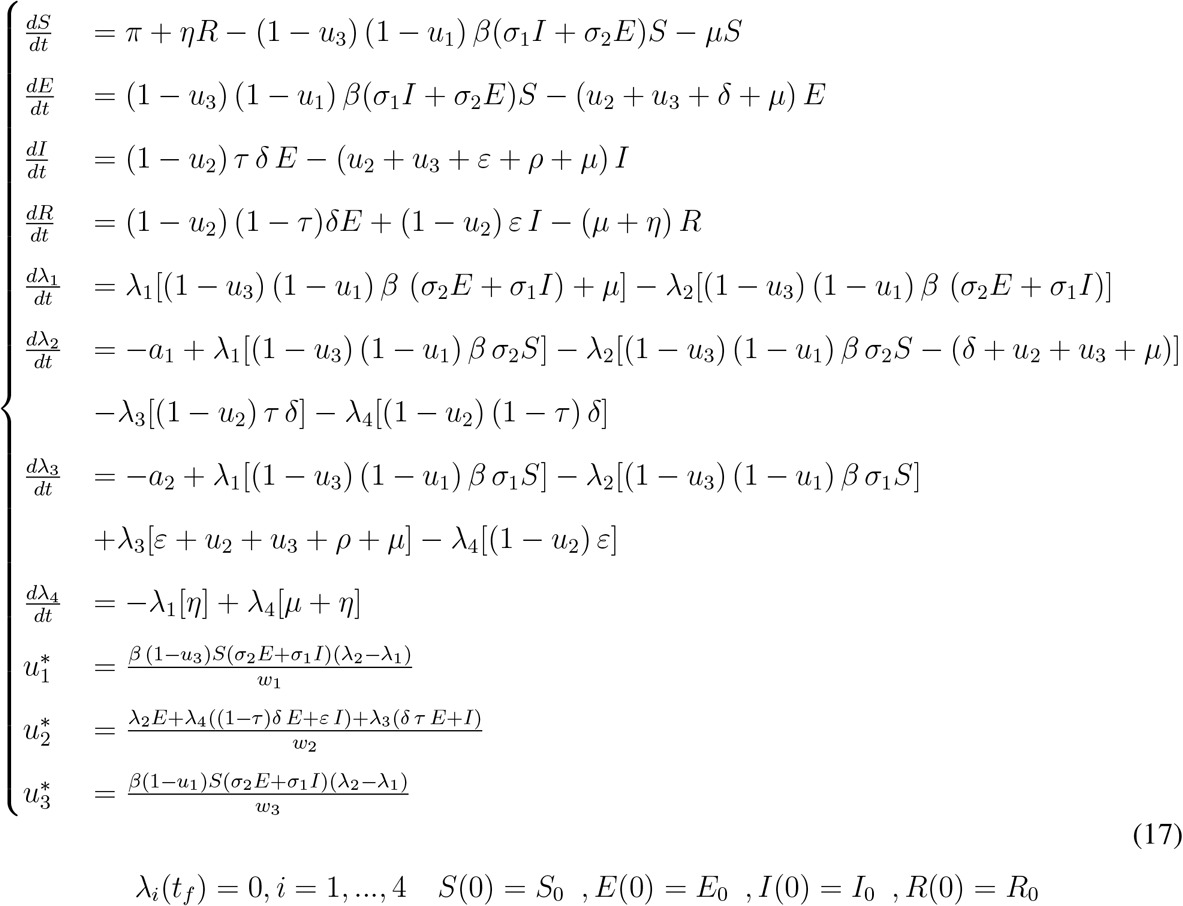

### 4.2 Uniqueness of the Optimality System

Due to the a priori boundedness of the state, adjoint functions and the resulting Lipschitz structure of the ODEs, we can obtain the uniqueness of solutions of the optimality system for the small time interval. Hence the following theorem

#### Theorem 4.2.

*For t ∈* [0, *t*_*f*_], *the bounded solutions to the optimality system are unique. For the proof of the theorem [26]*.

## 5 Numerical Simulations

In this section, we examine the COVID 19 model and studied the effects of combined strategies on controlling the transmission of the disease. The optimal control set was obtained by solving the optimality system, consisting of the state and adjoint systems. An iterative scheme was used for solving the optimality system. We start to solve the state equations with an initial guess for the controls over the simulated time using the forward fourth order Runge–Kutta scheme. Because of the transversality conditions, the adjoint equations were solved by a backward fourth order Runge–Kutta scheme using the current iterated solutions of the state equation. Then the controls are updated by using a convex combination of the previous controls and the value from the characterizations. This process is repeated and iterations are stopped if the values of the unknowns at the previous iteration are very close to the ones at the present iteration. For numerical simulation purpose, we have used empirical data of COVID-19 cases in Ethiopia for estimation of parameters in Table 3.

**Table 3:** Parameter values for COVID-19 model.

| Parameter symbol | Value | Source |
| --- | --- | --- |
| $\pi$ | 13.5 | Estimated |
| $\beta$ | 0.0143 | Estimated |
| $\sigma_1$ | 0.0001 | Assumed |
| $\sigma_2$ | 0.02 | Assumed |
| $\delta$ | 0.07 | Estimated |
| $\mu$ | 0.016 | Estimated |
| $\rho$ | 0.0004 | Estimated |
| $\tau$ | 0.7 | Assumed |
| $\epsilon$ | 0.15 | Estimated |
| $\eta$ | 0.15 | Estimated |

We investigated numerically the effect of the following optimal control strategies on the spread of COVID 19 in a population. Considering strategies that implement one intervention only is not guaranteed to reduce and/or eradicate the disease totally from the community. So that those strategies which incorporate more than one intervention are ordered below and compared pairwise:

- **Strategy A:** Preventive measures (*u*_1_) & supporting infectives with medication (*u*_2_)
- **Strategy B:** Preventive measures (*u*_1_) & media campaign (*u*_3_)
- **Strategy C:** Supporting infectives with medication (*u*_2_) & media campaign (*u*_3_)
- **Strategy D:** Using all control techniques (*u*_1_,*u*_2_ & *u*_3_)

In addition to those parameter values in Table 3, we used *a*_1_ = 1, *a*_2_ = 5, *w*_1_ = 40, *w*_2_ = 150 and *w*_3_ = 75 for simulation of COVID 19 pandemic disease model.

### 5.1 Strategy A: Control with preventive measures & supporting infectives with medication

Here we have investigated the impact of optimal prevention methods and supporting infected individuals in quarantine center with medication treatment. From the simulation results of Figure 2, we see that the combination of the two method is effective in controlling COVID-19 in the specified period. Moreover, we can see that after implementing this strategy the number of exposed and infected human population goes to zero after 15 months.

**Figure 2:**
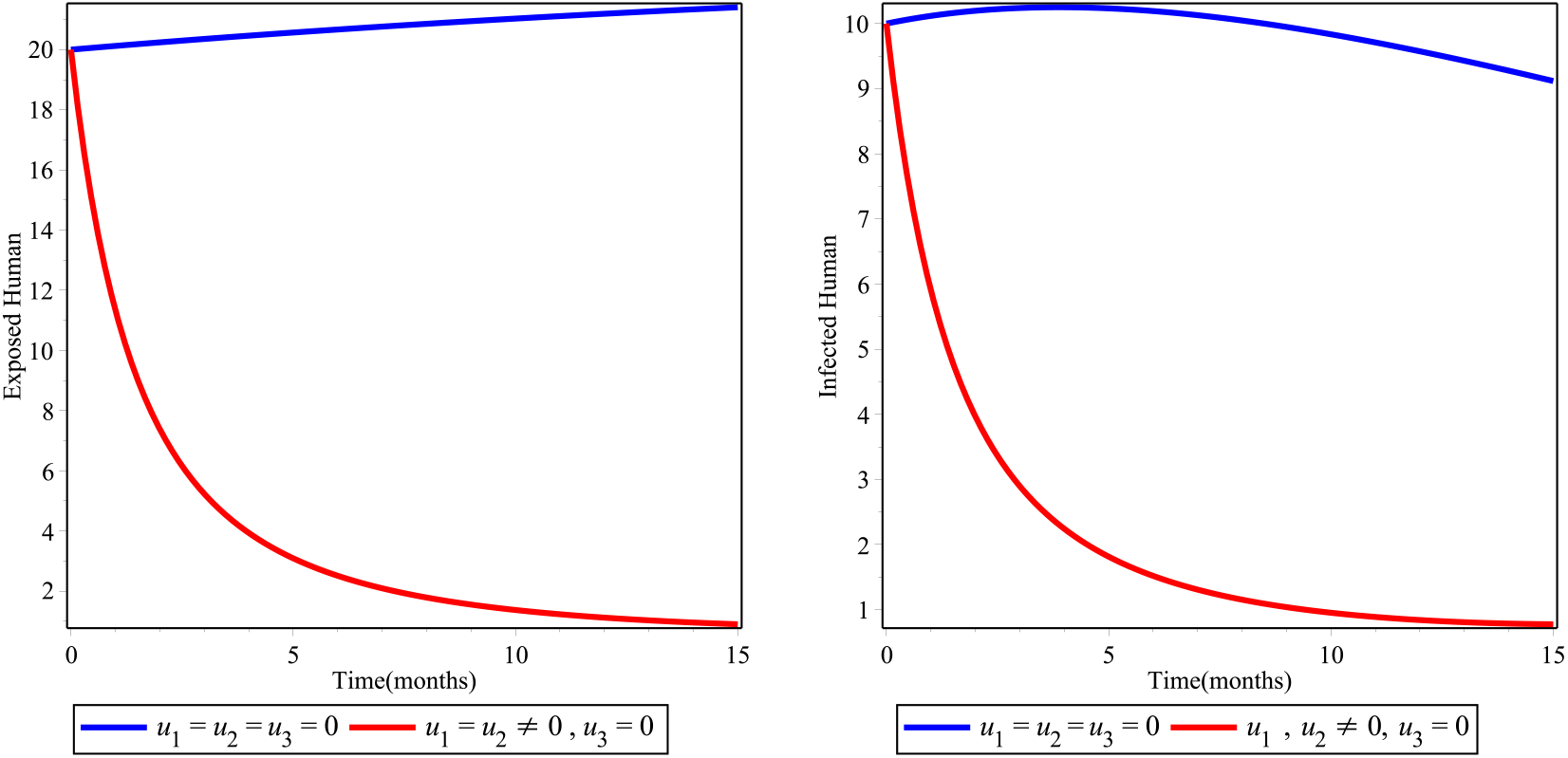
Simulations of the COVID 19 model with preventive measure & supporting infectives with medication.

### 5.2 Strategy B: Control with preventive measures & media campaign

Here also we experimented by combining preventive measure and awareness creation through media campaign. From the numerical result depicted in Figure 3 below (the left hand side) show as it is possible to combat the exposed human population of COVID-19 to zero after 8 months and it will also start to relapse again after 10 months. The right hand side of Figure 3 also indicate that as it is possible to minimize infected-COVID-19 humans but unable to eliminate by this strategy in the stated time. Also this method is not effective once the infectious individuals are entered in the country.

**Figure 3:**
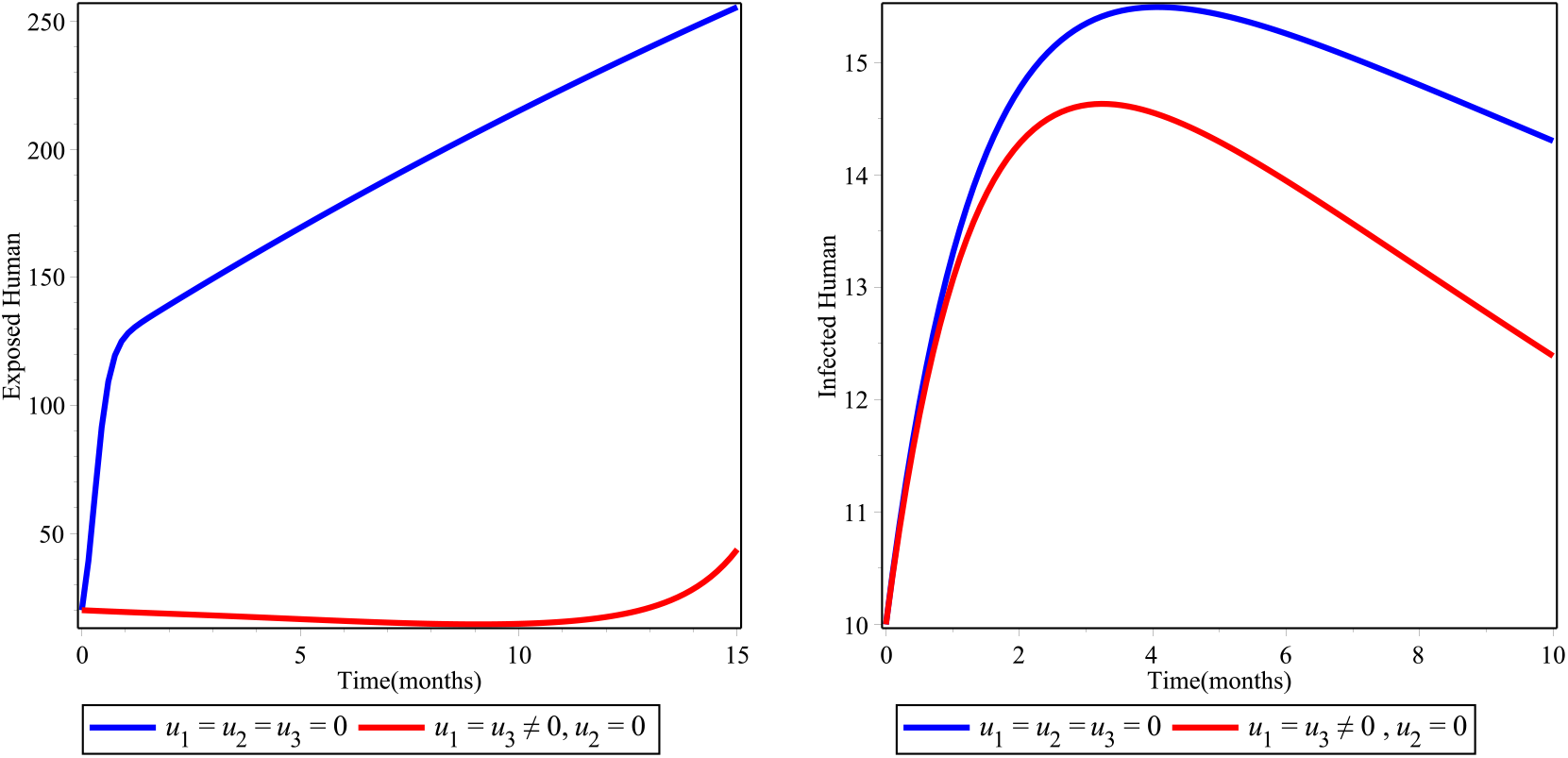
Simulations of the COVID 19 model with Social distancing with personal hygiene & Media campaign.

### 5.3 Strategy C: Control with supporting infectives by medication & media campaign

Here we experimented the model considering optimal support of infected individuals in quarantine center and creating awareness through media. Any activities from preventive technique of the disease is not considered here. From the numerical result depicted in Figure 4 shows that as COVID-19 will relapse for second time after it goes to zero in the specified time.

**Figure 4:**
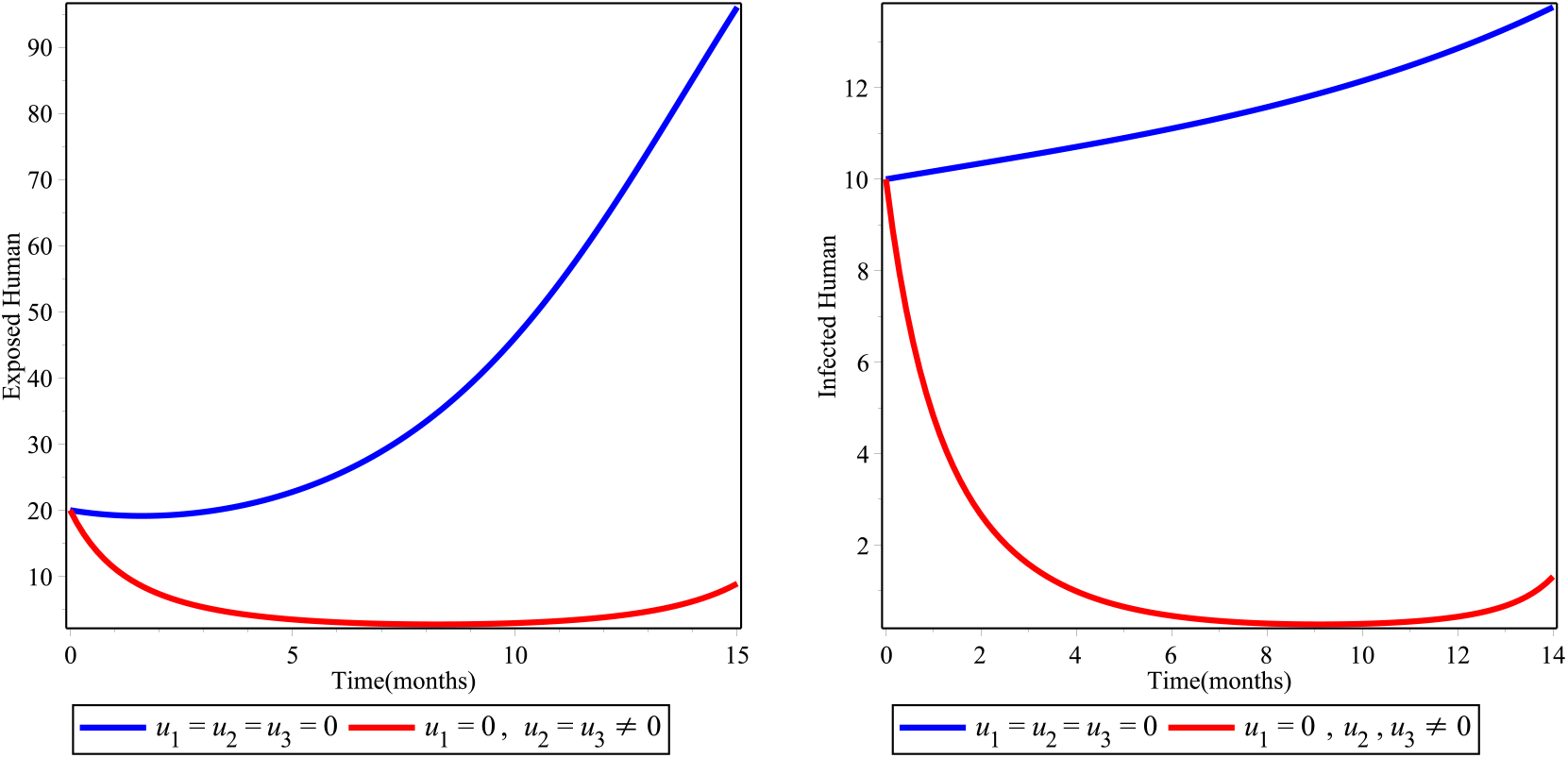
Simulations of the COVID 19 model with Supporting infectives with medication & Media campaign

### 5.4 Strategy D: Using All Control Strategies

Here we have applied allstrategies to optimlize the objective function. The results from Figure 5 shows that bringing down the exposed and infected population in short period of time and it is best compared to all the above three control technique. Therefore, government decision makers and all stakeholders consider in applying all strategies to combat COVID-19 in the specified time.

**Figure 5:**
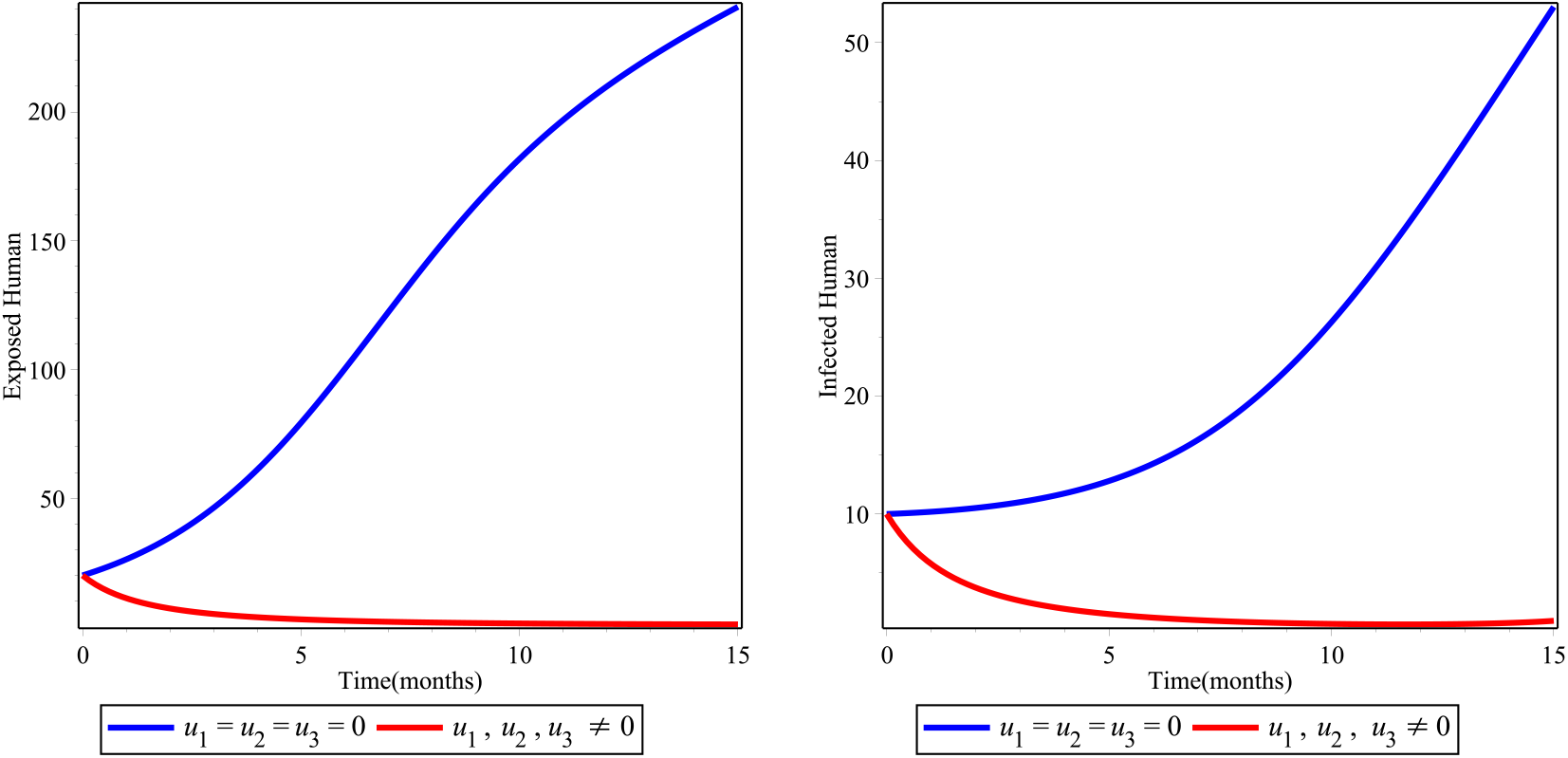
Simulations of the model with all control strategies.

## 6 Discussions and Conclusions

In this paper an SEIR deterministic model for the transmission dynamics of the pandemic COVID-19 in the case of Ethipia was formulated. Model analysis demonstrates that its solutions are positive and bounded, and there is a region where the model is well-posed mathematically and epidemiologically meaningful. The basic reproduction number ℜ_0_ was computed and the stability of equilibria points was investigated. Through Lyapunov’s theory, the disease free equilibrium point globally asymptotically stable whenever the ℜ_0_ < 1 was proven. Using center mainfold theory, bifurcation analysis of the model was proven and the model exhabts forward bifurication at ℜ_0_ = 1.

Second, by adding three times-dependent controls, we extend the basic model in to an optimal control. By using Pontryagin’s Maximum Principle necessary conditions for the optimal control of the transmission of COVID-19 were derived. From the numerical simulation it was found that the integrated control strategy, strategy D: using all technique, is very efficient in short period of time. Therefore, applying all rounded technique by the EFDRE, decision makers and stakeholders have a significant contribution in combating this pandemic in very short period of time. The result also shows, if the stakeholders could not do all of the intervention strategies together, the pandemic may come agian and will transmit over the country.

This paper is still an ongoing research as many more investigations regarding his disease can be carried out. Yet, it serves as the starting phase to research more in depth on questions that COVID-19 is spreading with incredible speed and have severe consequences. Moreover, to identify those exposed individuals who doesn’t develop clinical sign, mass screening in any cost is recommended to combat COVID-19.

## Data Availability

All data relevant to this publication will be provided when needed.

## Data Availability

All data relevant to this publication will be provided when needed.

## Conflicts of Interest

Authors have declared that no competing interests exist.

## Authors’ contribution

All authors contributed equally.

## Notes

### Competing Interest Statement

The authors have declared no competing interest.

### Clinical Protocols

http://www.hailegech.com

### Funding Statement

Not Applicable

### Author Declarations

Haileyesus Tessema Alemneh

